# The Effect of Zalunfiban on High Sensitivity Cardiac Troponin and the Association with Clinical Outcomes in Patients with STEMI

**DOI:** 10.64898/2026.02.03.26345522

**Authors:** James L. Januzzi, C. Michael Gibson, Gerald Chi, Barry S. Coller, Christopher B. Granger, Gilles Montalescot, Sem A.O.F. Rikken, Ashley Verberg, Jurrien M. Ten Berg, Arnoud W. J. van ‘t Hof, the CELEBRATE Investigators

**Author notes:** **Correspondence:** James L. Januzzi, Baim Institute for Clinical Research, 930 Commonwealth Avenue, Boston, MA, USA, E.

## Abstract

**Background:** Among patients with ST segment elevation myocardial infarction (STEMI), higher concentrations of high sensitivity troponin T (hs-cTnT) are associated with larger MI size and predict a worse prognosis. In the 2467 patient CELEBRATE trial, a single subcutaneous injection of the short-acting glycoprotein IIb/IIIa receptor blocker antagonist zalunfiban at first medical contact significantly improved the primary outcome including clinical endpoints. In this study, we assessed the impact of zalunfiban on MI size and association with downstream outcomes remains unclear.

**Methods:** In a prespecified analysis, we studied results among study participants treated with two doses of zalunfiban who had core laboratory measurements concentrations of hs-cTnT.

**Results:** The median concentration of hs-cTnT at presentation was 62 ng/L; at 24 hours it was 1962 ng/L. More elevated hs-cTnT concentrations at presentation were associated with less resolution of ST deviation after revascularization (P =0.006) and more frequent Q wave development (all P <0.001). At coronary angiography more elevated hs-cTnT at presentation was also associated with higher thrombus grade and worse epicardial and myocardial perfusion (all P <0.05). In multivariable analyses, higher hs-cTnT concentrations at 24 hours were associated with greater adjusted risk for all-cause death (odds ratio [OR] 1.83 per log unit increase; P=0.03), cardiovascular death (OR 1.83 per log unit increase; P=0.03), heart failure (OR 2.74 per log unit increase; P <0.001) or the composite of death (or cardiovascular death) and heart failure (P<0.001) by 30 days. At 24 hours, those treated with zalunfiban had lower hs-cTnT compared to placebo (1900 vs 2082 ng/L; P =0.04) and across multiples ≥10 to ≥1000 times elevation, treatment with zalunfiban resulted in smaller hs-cTnT determined MI size.

**Conclusion:** Among patients with STEMI, higher concentrations of hs-cTnT are associated with worse angiographic and ECG measures of reperfusion. More elevated hs-cTnT predicted a higher risk for short-term death or heart failure. A single dose of zalunfiban at first medical contact reduced MI size, as judged by more study participants with lower hs-cTnT concentrations. These results provide a mechanistic basis for the improved clinical outcomes associated with zalunfiban treatment in the CELEBRATE Trial.

**Study registration:** A Phase 3 Study of Zalunfiban in Subjects With ST-elevation MI (CELEBRATE); NCT04825743

---

ST-segment elevation myocardial infarction (STEMI) remains a leading cause of cardiovascular morbidity and mortality worldwide, with clinical outcomes strongly influenced by the extent of myocardial injury sustained before and during reperfusion therapy^1-6^. Cardiac troponin has emerged as a robust biomarker of infarct size, with higher concentrations consistently associated with larger MI, impaired myocardial recovery, and increased short-term and long-term adverse events^1-6^. These worse outcomes are largely related to worse measures of reperfusion, caused by platelet-rich thrombotic burden within and downstream to the infarct-related lesion^7^; greater thrombus burden directly results in worse epicardial and microvascular flow^8^, and in turn, larger MI sizes with worse outcome. Despite advances in systems of care and rapid reperfusion strategies, early thrombotic burden and microvascular obstruction continue to limit myocardial salvage in many patients.

Glycoprotein IIb/IIIa receptor blockade has been recognized as a potent antiplatelet strategy capable of reducing intracoronary thrombus formation, facilitating reperfusion, and improving coronary flow^9-12^, yet clinical use of glycoprotein IIb/IIIa receptor blockade in STEMI has been constrained by bleeding risk, thrombocytopenia, and the need for intravenous administration^13, 14^. Zalunfiban (RUC-4), a novel short-acting, subcutaneously administered glycoprotein IIb/IIIa inhibitor^14, 15^, offers a potentially transformative approach by enabling potent platelet inhibition at first medical contact without prolonged drug exposure owing to its very short half-life of one hour^16^. Zalunfiban also differs from current small molecule glycoprotein IIb/IIIa inhibitors in not inducing conformational changes in the receptor^17^; this may reduce risk for thrombocytopenia after zalunfiban treatment and thereby further mitigate risk of bleeding^14, 15^.

In the Phase 3 Study of Zalunfiban in Subjects With ST-elevation MI (CELEBRATE) trial^18, 19^, a single dose of zalunfiban prior to catheterization resulted in a significantly lower odds of a composite endpoint including death, stroke, recurrent MI, stent thrombosis, heart failure (HF) and larger MI size^19^. Zalunfiban also improved measures of reperfusion, including infarct artery patency and ST segment resolution on electrocardiography (ECG)^19^. Collectively, the results of the CELEBRATE trial suggest that early platelet inhibition enhances myocardial reperfusion and reduces downstream thrombotic complications. However, the mechanistic basis for these benefits—particularly the effect of zalunfiban on infarct size and its relationship to subsequent clinical events—has not been fully elucidated.

We conducted a prespecified analysis of troponin concentrations measured in participants enrolled in CELEBRATE. In this study, we compare biomarker-derived infarct size between patients treated with zalunfiban and those receiving placebo, and examine the association among troponin concentrations, angiographic characteristics, and short-term clinical outcomes to clarify the biological and clinical impact of early glycoprotein IIb/IIIa inhibition in STEMI. Understanding whether zalunfiban meaningfully reduces myocardial injury may provide critical insight into the mechanisms underlying its observed benefits and inform future strategies for pre-hospital management of STEMI.

## Methods

The CELEBRATE trial was designed and managed jointly by the Executive Committee of the study and the sponsor, CeleCor Therapeutics. The Executive Committee of the trial had full access to all the data in the study and takes responsibility for its integrity. The sponsor had no role in the data analysis, data interpretation, or writing of the manuscript. A medical writer was not used for the development of the manuscript.

The CELEBRATE protocol and all amendments were approved by the institutional review board or ethics committee at each participating site and consent was obtained according to local regulations before enrollment.

### Study Design and Population

The methods and primary results of the CELEBRATE trial have been previously reported^18, 19^. In brief, prospective adult trial participants were eligible if they had ischemic chest pain lasting more than 10 minutes and presented within 4 hours of symptom onset; they also were required to have ECG evidence of ST-segment elevation of at least 2 mm in contiguous leads. Exclusion criteria included cardiogenic shock, out-of-hospital cardiac arrest, current use of oral anticoagulation, renal replacement therapy, recent major bleeding, recent surgery, or a history of stroke.

Participants were enrolled at first medical contact and randomized to receive a single subcutaneous injection of zalunfiban or matching placebo prior to hospital arrival; the majority were consented during ambulance transport. Enrolled patients meeting all eligibility criteria and consented to enrollment were randomly assigned in a 1:1:1 ratio to receive either a placebo or zalunfiban at doses of 0.11 mg/kg or 0.13 mg/kg. Per protocol, blood samples were obtained for troponin measurement at presentation and 24±12 hours following presentation.

### Current Analysis

The present analysis was prespecified within the CELEBRATE statistical analysis plan. For analyses involving baseline high-sensitivity cardiac troponin T (hs-cTnT) results, those study participants with available core laboratory hs-cTnT results at presentation were included; for analyses involving the 24 hour time point, those with hs-cTnT at 24 hours were included. For the purposes of this analysis, both dosing arms of zalunfiban were pooled as was prespecified in the statistical plan for the trial.

### Biomarker Assessment

Concentrations of troponin were assessed using a validated 5^th^ generation hs-cTnT method (Roche Diagnostics, Mannheim, GE). The upper reference limit for this assay is 14 ng/L regardless of sex. Troponin measurement was performed in a core laboratory following study procedures (Department of Clinical Chemistry, Isala, Zwolle, NL).

### Clinical and Angiographic Outcomes

As reported, all clinical outcomes were categorized by an Event Adjudication Committee blinded to core laboratory hs-cTnT concentrations, and included all-cause death, cardiovascular (CV) death, heart failure (HF) events, and a composite of death (or CV death) or HF through 30 days. Angiographic characteristics—thrombus grade, epicardial flow, and myocardial perfusion—were assessed by similarly blinded core laboratory readers (Perfuse, Boston, MA). ECG endpoints included ST-segment deviation resolution before and after revascularization and Q-wave development after revascularization; ECG findings were assessed in a blinded manner by an independent core laboratory (Diagram, Zwolle, NL).

## Statistical Analysis

Continuous variables were summarized using means and standard deviation (SD) or medians and first/third quartile (Q1, Q3), depending on normality. Categorical and continuous variables were compared with the chi-square and Kruskal-Wallis test, respectively. Associations between hs-cTnT concentrations and clinical outcomes by 30 days were evaluated using logistic regression; models adjusted for relevant clinical covariates including age, male sex, time from symptom onset to study drug, anterior MI location and kidney function (estimated glomerular filtration rate). For clinical outcomes modeling, concentrations of hs-cTnT were log2-transformed, with odds ratios (ORs) reported per log-unit increase in log2-hs-cTnT; ORs were generated with 95% confidence intervals (CI). Concentrations of hs-cTnT were compared between treatment groups at baseline and 24 hours and a cumulative distribution plot evaluated distribution of hs-cTnT at 24 hours among study participants treated with zalunfiban versus placebo, and expressed relative to multiples of the upper hs-cTnT reference limit.

All statistics were performed SAS® version 9.4 (Cary, NC). P values are two-sided with <0.05 considered statistically significant.

## Results

### Study Population

From the total sample of 2467 participants in the CELEBRATE trial, 2172 had hs-cTnT results at baseline (1473 were treated with zalunfiban and 699 were treated with placebo). At 24 hours post PCI, 1758 had hs-cTnT results (1186 were treated with zalunfiban and 572 were treated with placebo). Baseline characteristics for both samples are compared to the overall CELEBRATE population in **Supplemental Tables 1 and 2**, which show no major differences. The study participants in the present analysis had a mean (±SD) age of 63 ± 11.2 years and approximately 80% were male. Greater than >90% of study participants underwent percutaneous coronary intervention (PCI) and nearly all received a P2Y12 inhibitor (mostly ticagrelor) and heparin at the same time that they received study drug or placebo.

### Baseline hs-cTnT

Among those study participants with baseline hs-cTnT values, the median (Q1, Q3) concentration at presentation was 62 (27, 165) ng/L; those treated with zalunfiban had a baseline hs-cTnT of 61 (26, 165) ng/L, while those treated with placebo had a baseline hs-cTnT of 63 (29, 161) ng/L.

Characteristics of study participants as a function of baseline hs-cTnT quartiles are detailed in **Table 1**. This shows that those with more elevated hs-cTnT concentrations at baseline tended to have a higher risk profile, with more advanced age, worse kidney function, more prolonged time from ischemic symptom onset and longer time to treatment (all P <0.002). They were also more likely to have higher heart rates and blood pressure, and to have ECG evidence for anterior wall MI (all P <0.001). Those with higher hs-cTnT concentrations also had the highest concentrations of N-terminal pro-B type natriuretic peptide (NT-proBNP; P <0.001).

**Table 1:** Characteristics of study participants as a function of hs-cTnT at study entry. The P-value refers to the trend across quartiles.

|  | **hs-cTnT Q1**<br>**(N=544)** | **hs-cTnT Q2**<br>**(N=542)** | **hs-cTnT Q3**<br>**(N=543)** | **hs-cTnT Q4**<br>**(N=543)** | **P-value** |
| --- | --- | --- | --- | --- | --- |
| **Age, years, mean (SD)** | 61.5 (10.5) | 63.1 (11.1) | 63.3 (11.5) | 64.3 (11.7) | 0.001 |
| **BMI, kg/m2, mean (SD)** | 27.6 (4.7) | 27.5 (4.4) | 27.3 (4.1) | 27.3 (4.3) | 0.72 |
| **Male, n (%)** | 446 (82.0%) | 439 (81.0%) | 418 (77.0%) | 428 (78.8%) | 0.17 |
| **White, n (%)** | 520 (95.6%) | 515 (95.0%) | 519 (95.6%) | 526 (96.9%) | 0.48 |
| **Current smoker, n (%)** | 232 (42.8%) | 203 (37.7%) | 222 (41.3%) | 235 (43.4%) | 0.22 |
| **Diabetes, n (%)** | 84 (15.4%) | 72 (13.3%) | 79 (14.5%) | 97 (17.9%) | 0.20 |
| **Hypercholesterolemia, n (%)** | 72 (13.2%) | 80 (14.8%) | 75 (13.8%) | 69 (12.7%) | 0.79 |
| **Hypertension, n (%)** | 172 (31.6%) | 197 (36.3%) | 186 (34.3%) | 214 (39.4%) | 0.05 |
| **Previous MI, n (%)** | 64 (11.8%) | 53 (9.8%) | 20 (3.7%) | 27 (5.0%) | <.001 |
| **Symptom onset to diagnostic ECG, minutes, median (Q1, Q3)** | 46.0 (27.0, 80.0) | 61.0 (33.0, 109.0) | 83.0 (44.0, 148.0) | 113.0 (56.0, 185.0) | <.001 |
| **Symptom onset to study drug, minutes, median (Q1, Q3)** | 65.0 (45.0, 101.0) | 80.0 (52.0, 134.0) | 105.0 (64.0, 175.0) | 138.0 (80.0, 212.0) | <.001 |
| **Study drug to PCI, minutes, median (Q1, Q3)** | 45.0 (36.0, 58.0) | 48.0 (39.0, 60.0) | 51.0 (41.0, 62.0) | 54.0 (43.0, 68.0) | <.001 |
| **Heart rate, beats/minute mean (SD)** | 69.3 (15.2) | 74.1 (16.6) | 76.9 (15.9) | 82.1 (17.4) | <.001 |
| **Systolic BP, mm Hg, mean (SD)** | 137.0 (24.9) | 143.9 (25.6) | 147.3 (25.6) | 148.4 (24.8) | <.001 |
| **Diastolic BP, mm Hg, mean (SD)** | 82.8 (16.3) | 86.9 (16.7) | 88.9 (16.9) | 90.4 (16.3) | <.001 |
| **Anterior MI, n (%)** | 152 (28.9%) | 201 (38.7%) | 221 (41.3%) | 276 (51.9%) | <.001 |
| **Antithrombotic pre-treatment, n (%)** |  |  |  |  | 0.10 |
| Heparin + P2Y12i | 513 (94.3%) | 519 (95.8%) | 519 (95.6%) | 522 (96.1%) |  |
| Heparin only | 9 (1.7%) | 1 (0.2%) | 3 (0.6%) | 2 (0.4%) |  |
| No heparin or P2Y12i | 22 (4.0%) | 22 (4.1%) | 21 (3.9%) | 19 (3.5%) |  |
| **eGFR, mL/min/1.73m2, mean (SD)** | 85.0 (13.0) | 81.9 (14.4) | 82.0 (15.9) | 77.6 (18.6) | 0.004 |
| **NT-proBNP, ng/L, median (Q1, Q3)** | 786 (396, 1340) | 1095 (618, 2180) | 1325 (715, 2485) | 2050 (1120, 4080) | <.001 |

SD denotes: standard deviation; BMI denotes: body-mass index; MI denotes: myocardial infarction; ECG denotes: electrocardiogram;
Q denotes: quartile; PCI denotes: percutaneous coronary intervention; BP denotes: blood pressure; mmHg denotes: millimeters of
mercury; eGFR denotes: estimated glomerular filtration rate; mL/min/1.73m2 denotes: milliliters per minute per 1.73 squared meters;
NT-proBNP denotes: N-terminal pro-B type natriuretic peptide; ng/L denotes: nanograms per liter

Results of ECG and coronary angiography and as a function of baseline hs-cTnT quartiles are detailed in **Table 2 and Table 3**. On ECG, those with higher concentrations of hs-cTnT at presentation had more extreme ST segment deviation compared to lower hs-cTnT quartiles (P <0.001). Those with higher concentrations showed a trend toward lower likelihood to have resolution of ST elevation prior to PCI (P = 0.07 across quartiles), and were significantly less likely to have ST segment resolution 1 hour post-PCI (P =0.006) compared to those with lower hs-cTnT concentration. Those with higher presenting hs-cTnT concentrations were also considerably more likely to develop pathological Q waves by 72 hours post PCI (or by hospital discharge; P <0.001).

**Table 2:** Results of electrocardiography findings as a function of hs-cTnT at baseline. The P-value refers to the trend across quartiles.

|  | **hs-cTnT Q1**<br>**(N=544)** | **hs-cTnT Q2**<br>**(N=542)** | **hs-cTnT Q3**<br>**(N=543)** | **hs-cTnT Q4**<br>**(N=543)** | **P-value** |
| --- | --- | --- | --- | --- | --- |
| **ST deviation, baseline, mm, median (Q1, Q3)** | 13.1 (8.6) | 14.3 (8.8) | 14.0 (7.6) | 14.8 (8.2) | 0.001 |
| **ST deviation resolution, pre-PCI, %, median (Q1, Q3)** | 16.1 (-16.7, 47.1) | 18.4 (-11.1, 44.4) | 21.8 (-6.0, 49.2) | 14.3 (-7.7, 40.0) | 0.07 |
| **ST deviation resolution, 1 hour post-PCI, %, median (Q1, Q3)** | 66.7 (45.0, 84.2) | 68.8 (47.1, 83.3) | 68.5 (44.8, 83.6) | 63.6 (40.7, 78.6) | 0.006 |
| **Pathologic Q waves, 72 hours post-PCI/hospital discharge, n (%)** | 308 (60.3%) | 336 (65.0%) | 356 (69.5%) | 407 (78.4%) | <.001 |

Q denotes: quartile; PCI denotes: percutaneous coronary intervention

**Table 3:** Results of coronary angiography findings as a function of hs-cTnT at baseline. The P-value refers to the trend across quartiles.

|  | **hs-cTnT Q1**<br>**(N=544)** | **hs-cTnT Q2**<br>**(N=542)** | **hs-cTnT Q3**<br>**(N=543)** | **hs-cTnT Q4**<br>**(N=543)** | **P-**<br>**value** |
| --- | --- | --- | --- | --- | --- |
| **Corrected TIMI frame count, pre-PCI, median (Q1, Q3)** | 176.0 (46.0,<br>176.0) | 170.5 (38.0,<br>176.0) | 99.0 (36.0,<br>176.0) | 83.5 (33.0,<br>176.0) | <.001 |
| **TIMI grade 3 myocardium perfusion, pre-PCI, n (%)** | 126 (25.4%) | 158 (31.0%) | 166 (31.9%) | 172 (33.3%) | 0.03 |
| **TIMI thrombus grade, pre-PCI, n (%)** |  |  |  |  | 0.04 |
| 0 | 67 (13.4%) | 83 (16.3%) | 86 (16.4%) | 85 (16.4%) |  |
| 1 | 63 (12.6%) | 67 (13.1%) | 81 (15.5%) | 93 (18.0%) |  |
| 2 | 11 (2.2%) | 11 (2.2%) | 18 (3.4%) | 10 (1.9%) |  |
| 3 | 46 (9.2%) | 50 (9.8%) | 58 (11.1%) | 51 (9.8%) |  |
| 4 | 33 (6.6%) | 55 (10.8%) | 48 (9.2%) | 40 (7.7%) |  |
| 5 | 279 (55.9%) | 244 (47.8%) | 232 (44.4%) | 239 (46.1%) |  |
| **TIMI grade 3 flow, 1-hour post-PCI, n (%)** | 436 (89.2%) | 443 (89.1%) | 428 (85.4%) | 397 (81.0%) | <.001 |
| **Corrected TIMI frame count, 1-hour post-PCI, median (Q1, Q3)** | 22.0 (15.0, 30.0) | 22.0 (16.0, 30.0) | 23.0 (18.0, 32.0) | 24.0 (18.0, 36.0) | 0.003 |
| **TIMI grade 3 myocardium perfusion, 1-hour post-PCI, n (%)** | 269 (55.8%) | 277 (55.8%) | 259 (52.1%) | 218 (45.4%) | 0.003 |

TIMI denotes: Thrombolysis in Myocardial Infarction; Q denotes: quartile; PCI denotes: percutaneous coronary intervention

Prior to PCI, across troponin quartiles, those with more elevated concentration of hs-cTnT at presentation were more likely to show higher intracoronary thrombus burden (P = 0.04) and they were more likely to have worse epicardial coronary flow, reflected in higher corrected Thrombolysis In Myocardial Infarction (TIMI) frame count (P <0.001); those with higher presentation hs-cTnT were also more likely to have worse corrected TIMI frame count following PCI (P <0.001) and were less likely to have TIMI grade 3 myocardial perfusion grade 1 hour post-PCI (P <0.001).

### Troponin Concentrations and Prognosis

From the baseline median concentration of 62 (27, 165) ng/L, the median concentrations of hs-cTnT rose to 1962 (788, 3900) by 24 hours. Across quartiles of hs-cTnT the rates of adjudicated death, CV death, HF, or composites of death/HF, CV death/recurrent MI, CV death/HF, or CV death/recurrent MI/HF within 30 days were all highest in those with more elevated hs-cTnT by 24 hours (**Figure 1**; all P <0.001).

**Figure 1.**
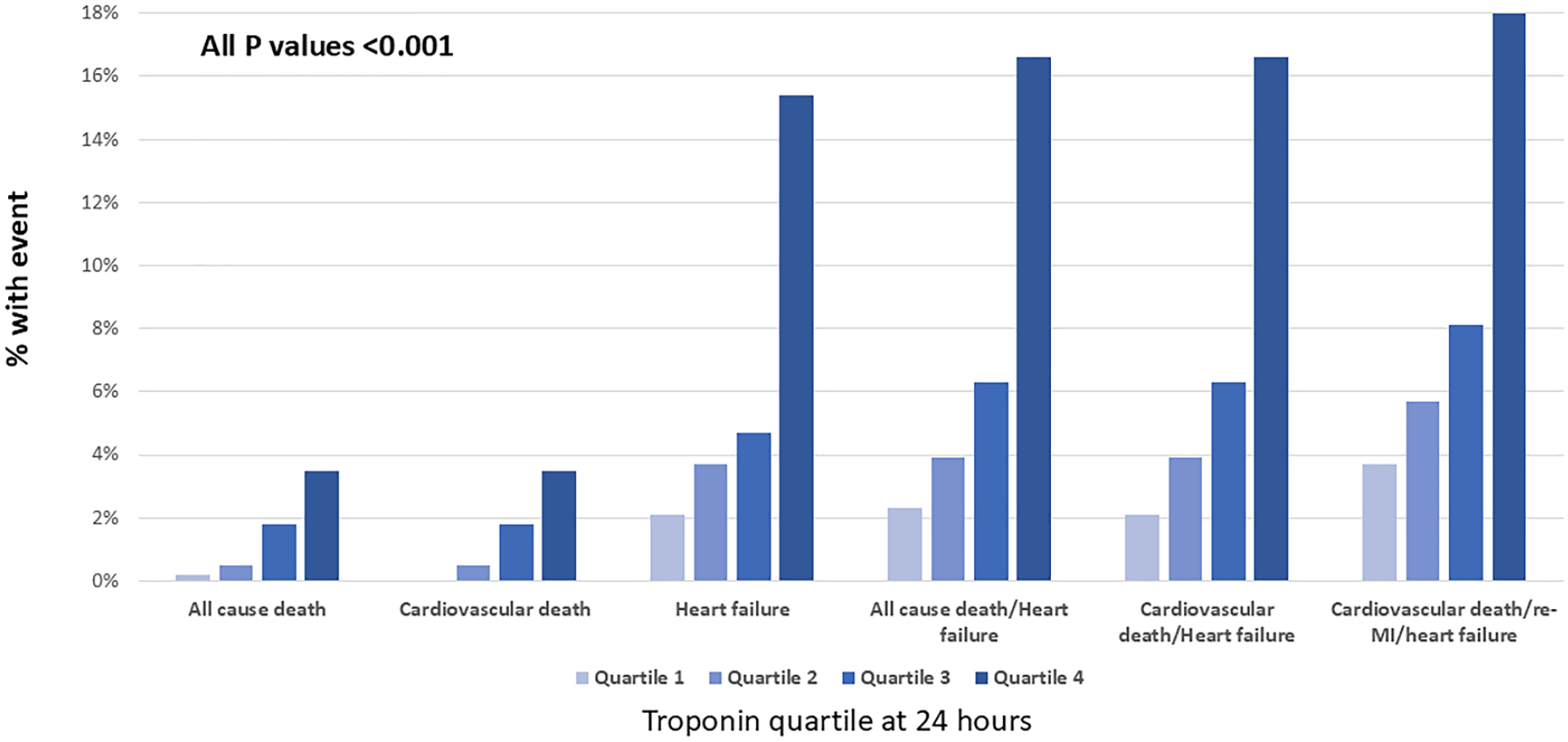
Cardiovascular events by 30 days after ST segment elevation myocardial infarction, expressed in quartiles of high sensitivity cardiac troponin T (hs-cTnT) at 24 hours after study enrollment. Across quartiles of hs-cTnT, higher frequencies of adverse outcome were noted.

In multivariable adjusted logistic regression for events by 30 days more elevated log2-hs-cTnT concentrations at 24 hours were independently predictive of all-cause death (OR = 1.83 per log unit increase, 95% CI = 1.06-3.17; P =0.03), CV death (OR = 1.83 per log unit increase, 95% CI = 1.06-3.17; P =0.03), HF (OR = 2.74 per log unit increase, 95% CI = 1.89-3.96; P <0.001), or the composite endpoints of all-cause death/HF (OR = 2.65 per log unit increase, 95% CI = 1.88-3.74; P <0.001), CV death/HF (OR = 2.65 per log unit increase, 95% CI = 1.88-3.74; P <0.001), or CV death/recurrent MI/HF (OR = 1.86 per log unit increase, 95% CI = 1.46-2.39; P <0.001).

### Effect of Zalunfiban on hs-cTnT Concentrations

Those treated with zalunfiban had lower hs-cTnT at 24 hours compared to placebo (1900 [740, 3800] vs 2082 [942, 4155] ng/L; P =0.04); participants treated with zalunfiban were also significantly less likely to have hs-cTnT concentrations ≥30× URL (P=0.04). Finally, as shown in the cumulative distribution function graph of hs-cTnT at 24 hours (**Figure 2**) treatment with zalunfiban resulted in a higher percentage of study participants with lower hs-cTnT concentrations at 24 hours across a broad range of values up to >1000X ULN a finding that is particularly noteworthy for hs-cTnT concentrations <500X ULN.

**Figure 2.**
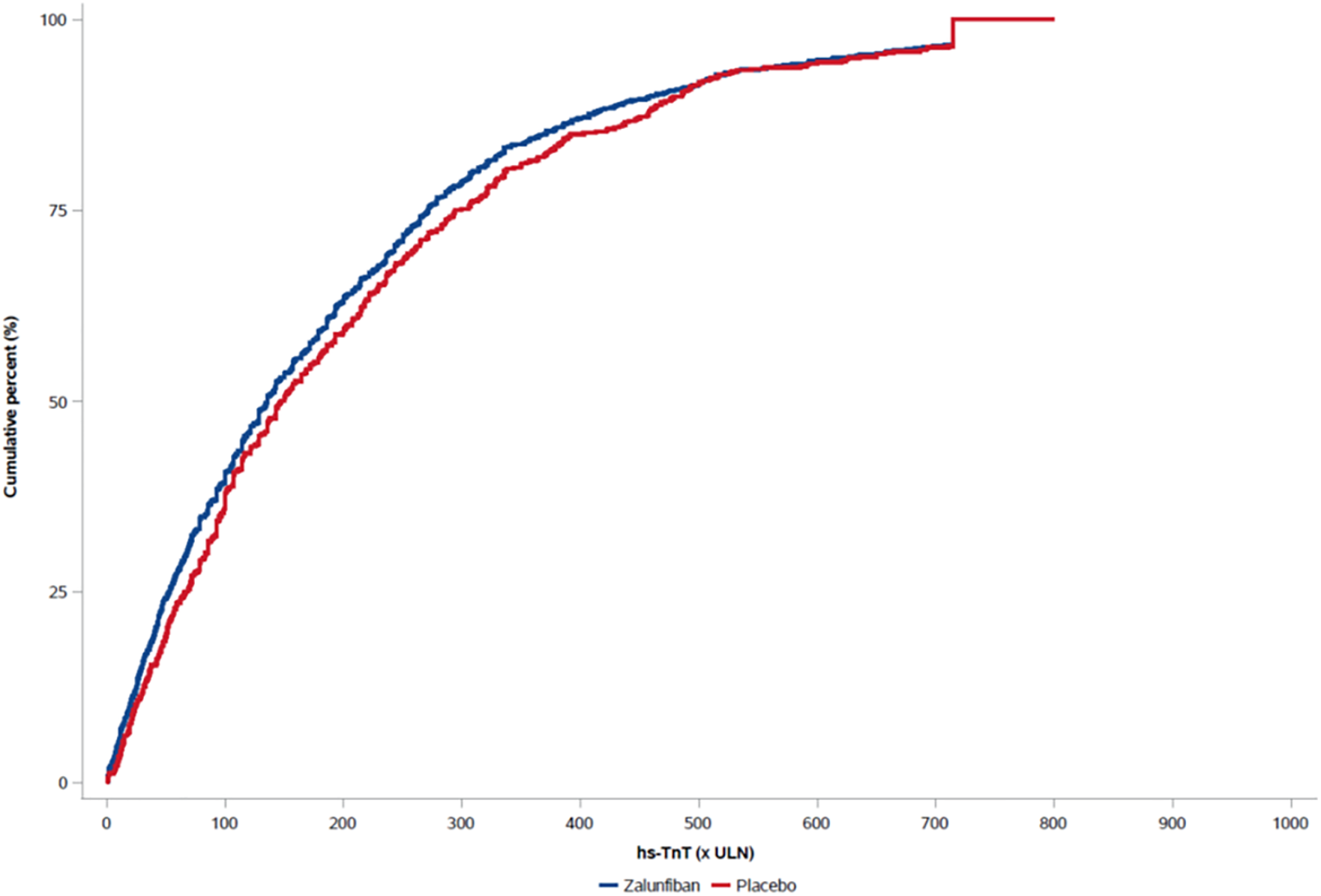
Cumulative distribution plot of hs-cTnT concentrations by 24 hours after study entry, expressed in multiples of the upper limit of normal (ULN). Results are depicted for those treated with zalunfiban (blue) and placebo (red). Across 0-1000X ULN, more study participants treated with zalunfiban had smaller MI size reflected in lower hs-cTnT concentration, a finding particularly noteworthy for hs-cTnT concentrations below 500X ULN.

## Discussion

In this prespecified analysis of the CELEBRATE trial, we observed several key findings. First, concentrations of hs-cTnT at presentation with STEMI were associated with higher-risk clinical features and worse angiographic findings. Second, more elevated hs-cTnT at 24 hours following STEMI was associated with greater adjusted risk for major adverse CV events, including mortality and particularly HF events during short-term follow-up. Third, early administration of the short-acting glycoprotein IIb/IIIa inhibitor zalunfiban at first medical contact was associated with smaller biomarker-estimated infarct size, as reflected by lower hs-cTnT concentrations at 24 hours. The ability of this ultra short-acting glycoprotein IIb/IIIa receptor antagonist to foster reperfusion prior to PCI and improve angiographic findings during and after PCI resulted in reduced MI size, and in turn, expected improved prognosis. This underscores the importance of very early platelet inhibition in STEMI care and provides important insights to the results of the CELEBRATE trial.

Troponin concentrations have a role as an indirect measure of MI size^1-5, 20-22^ but mechanistic correlates to explain such an association are less well understood. The present analysis considerably extends understanding of how concentrations of hs-cTnT link to MI size; in this analysis, baseline hs-cTnT values were associated with higher thrombus burden at coronary angiography, and were strongly associated with worse epicardial and microvascular perfusion, the latter of which largely determined by distal embolization, a major driver of infarct size^10^. In turn, higher troponin concentrations were correlated with impaired ST-segment resolution and a higher frequency of Q wave formation. It is noteworthy that treatment of STEMI with zalunfiban reduced intracoronary thrombus and improved epicardial and myocardial perfusion before angiography^19^; this is consistent with the reduced hs-cTnT concentrations associated with zalunfiban use.

Notably, more elevated hs-cTnT in those with STEMI was also associated with significantly higher adjusted risk for adverse clinical outcomes including death and HF. Higher troponin was also associated with more marked elevation of NT-proBNP, itself also associated with microvascular dysfunction in STEMI^23^ and predictive of worse outcomes (including death and HF) after MI^24^. Conceptually, greater epicardial and microvascular thrombus burden is associated with worse microvascular obstruction, greater transmurality of injury, and larger infarction size, reflected in higher hs-cTnT; this, in turn results in greater wall stress (reflected in more elevated NT-proBNP), a propensity toward adverse ventricular remodeling, and a worse prognosis^4^. These associations help reinforce the utility of troponin concentrations an integrated measure of infarct size and reperfusion quality: with contemporary revascularization strategies, the risk for major CV events following STEMI continues to decrease; it is thus important to have simple, widely available tools to identify those individuals at highest risk for poor outcomes. This includes identifying those most prone to early development of left ventricular dysfunction during follow up as such individuals may benefit from earlier application of guideline-directed medical therapy to prevent early myocardial remodeling and prevent early HF events or death^25^.

Post hoc results from trials of individuals with MI treated with tirofiban suggested glycoprotein IIb/IIIa receptor blockade might lower troponin concentrations compared to placebo^26, 27^, implying reduced MI size in this context. However, these post hoc studies were small and underpowered, and lacked the detailed clinical and angiographic data in the present prospective analysis. Furthermore, whether such results applied to those treated with more potent oral antiplatelet regimens and contemporary revascularization strategies remained unknown. In the present trial treatment with zalunfiban reduced MI size, with a more study participants showing MI sizes <500X ULN. This finding has therapeutic implications; greater epicardial burden of platelet-rich thrombus is associated with worse epicardial and myocardial flow^7, 8^, reflected in higher concentration of hs-cTnT as was shown in this study. Potent anti-platalet therapy with glycoprotein IIb/IIIa inhibitors may be particularly beneficial to reduce improve microvascular perfusion ^10^. The results of this analysis showing zalunfiban reduces MI size in parallel with improved measures of reperfusion and coronary flow strongly supports this hypothesis. Moreover, since lower 24 hour hs-cTnT concentrations were associated with reduced risk for mortality and HF events, these results highlight the clinical relevance of the reductions in myocardial injury produced by zalunfiban treatment.

## Limitations

Although this analysis is among the largest to evaluate the impact of glycoprotein IIb/IIIa receptor blockade on hs-cTn concentrations and link such an effect with clinical outcomes, certain considerations temper the interpretation of these results. Troponin release is influenced by multiple factors—including kidney function, ischemic duration, collateral flow, infarct territory, and reperfusion quality—making it challenging to isolate the effect of glycoprotein IIb/IIIa therapy^22^. Nonetheless, the results from this and other studies have emphasized importance of post-MI troponin concentrations as a key mechanistic outcome and a potential target for therapeutic intervention. Second, although hs-cTnT was predictive of mortality and HF events, given the expected low event rates for these outcomes, the CELEBRATE trial was not powered to show a statistically significant impact of zalunfiban on such hard CV outcomes. Nonetheless, zalunfiban did lower hs-cTnT, which in turn is associated with an expected reduction in harder CV outcomes. In this regard, our results point towards the potential importance of post-MI troponin concentrations as a key outcome measure in clinical trials of STEMI.

In conclusion, among individuals with STEMI, higher concentrations of hs-cTnT were associated with worse measures of early reperfusion related to a greater epicardial and microvascular thrombotic load. More elevated troponin was also linked to less ST segment resolution and more frequent Q wave formation. At 24 hours following STEMI, more elevated hs-cTnT was independently associated with mortality and HF events by 30 days in adjusted analyses. Notably, treatment with the ultra-short acting subcutaneous glycoprotein IIb/IIIa receptor blocker effectively lowered troponin concentrations. Early, potent, and transient glycoprotein IIb/IIIa inhibition with zalunfiban presumably enhanced myocardial salvage by reducing thrombus propagation and improving microvascular flow during the critical pre-reperfusion period. In aggregate, these results help to explain how zalunfiban treatment improved clinical outcomes in the CELEBRATE trial.

## Data Availability

Data will be available upon reasonable request.

## Non-standard Abbreviations and Acronyms

CELEBRATE: A Phase 3 Study of Zalunfiban in Subjects With ST-elevation MI
hs-cTnT: High sensitivity cardiac troponin T
CV: Cardiovascular
HF: Heart failure
TIMI: Thrombolysis in Myocardial Infarction

